# Unapproved Medicine Use by Paramedics in New Zealand: A Comparative Analysis with Australian and UK Frameworks

**DOI:** 10.1101/2024.11.10.24317076

**Authors:** Dylan A Mordaunt

## Abstract

**Objective:** To evaluate the regulation of unapproved medicines and its impact on paramedic practice in out-of-hospital settings by comparing regulatory frameworks in New Zealand, the United Kingdom, and Australia. The objective was to propose actionable policy recommendations to improve New Zealand’s current regulatory approach.

**Methods:** A comparative analysis was conducted using theoretical frameworks including regulatory theory, public health law, institutionalism, comparative policy analysis, and health crisis management. A technical comparison was also undertaken. Data were collected from legislative texts, policy documents, and secondary sources. The analysis focused on prescribing and administration authority, administrative requirements, flexibility in emergency situations, and the impact on patient care.

**Results:** New Zealand’s Section 29 of the Medicines Act 1981 imposes comprehensive reporting requirements and restricts unapproved medicine use to registered medical practitioners, hindering timely interventions by paramedics. The administrative burden and lack of flexibility in emergency situations compromise patient care. In contrast, the UK’s Human Medicines Regulations 2012 and Australia’s Therapeutic Goods Act 1989 provide structured and adaptable pathways. The Therapeutic Products Act 2023 in New Zealand proposed reforms but is currently in the process of being repealed.

**Discussion:** New Zealand’s framework of Section 29 is ill-suited for pre-hospital emergency care, creating ethical and practical dilemmas for paramedics. Comparative insights reveal that more flexible legal frameworks in the UK and Australia better support paramedics’ ability to provide timely care. Ethical considerations emphasise the need to balance regulatory oversight with patient care imperatives.

**Conclusions:** Legislative reforms in New Zealand are urgently needed to enable the lawful administration of unapproved medicines by paramedics, reduce administrative burdens, and align its framework with international best practices.

## Introduction

In out-of-hospital (urgent and emergency ambulance) care, paramedics are tasked with delivering life-saving interventions under intense time pressure, usually in isolation and often in challenging (austere) environments. Their ability to provide timely and effective care depends on the availability of appropriate medications. However, legal frameworks regulating medicine use can impose significant restrictions, particularly when paramedics need to administer unapproved medicines. In 2024 paramedics frequently need to administer unapproved medicines due to global supply chain disruptions ^1, 2^.

In New Zealand, Section 29 of the Medicines Act 1981 mandates that only registered medical practitioners may prescribe or administer unapproved medicines, and each instance must be reported to the Ministry of Health ^3^. While intended to maintain oversight, this provision creates practical challenges for paramedics who often operate independently and under considerable time constraints. The rigid nature of Section 29 contrasts with more flexible frameworks in Australia and the United Kingdom, which empower paramedics under defined emergency protocols ^4, 5^.

Since 2020, medicine supply chains have encountered significant disruptions due to a combination of global events and challenges ^1, 2^. The COVID-19 pandemic played a central role by causing widespread lockdowns, which led to reduced manufacturing output and labour shortages in production facilities worldwide ^1, 2^. Transportation restrictions and border closures further hindered the movement of raw materials and finished pharmaceutical products ^1, 2^. Additionally, the surge in demand for certain medications, personal protective equipment, and vaccines strained existing supply capacities ^1^. Geopolitical tensions, including trade disputes and export controls, have also impacted the availability of medicines by affecting international collaborations and supply agreements ^6^. Natural disasters and climate-related events have disrupted logistics and damaged infrastructure essential for the production and distribution of pharmaceuticals ^7^. Collectively, these factors have exposed vulnerabilities in the global medicine supply chain ^2^. In New Zealand, this is exacerbated further by the impact of our public payer model on the domestic medicines market and related supply chain resilience.

The realities of out-of-hospital emergency care differ substantially from clinical settings. Paramedics must make rapid decisions without the luxury of time or consultation, aiming to stabilise critically ill patients. Supply chain disruptions, manufacturing shortages, and public health emergencies like the COVID-19 pandemic have underscored the importance of flexible legal provisions that allow healthcare professionals to adapt quickly ^1, 2^. This article examines the limitations of Section 29 in New Zealand’s ambulance environment and explores potential reforms inspired by practices in Australia and the UK to enhance paramedics’ ability to deliver timely and effective care. The objective was to compare unapproved medicines regulatory frameworks and to propose actionable policy recommendations to improve New Zealand’s current regulatory approach

## Methods

### Methodological Approach

A comparative analysis was conducted to evaluate the regulatory frameworks governing the use of unapproved medicines in New Zealand, Australia, and the United Kingdom. These countries were selected due to similarities between the health systems. This methodology is appropriate for exploring the nuances of different legal systems and their impact on healthcare delivery, particularly in emergency settings ^8^.

This analysis is guided by five theoretical frameworks. Regulatory Theory examines how regulations are structured and enforced to balance control and flexibility ^6^. It considers the effectiveness of legal frameworks in achieving regulatory objectives without imposing unnecessary burdens on practitioners. Public Health Law evaluates the implications of laws on health outcomes, emphasising patient safety and accessibility ^9^. It assesses how legal provisions impact the delivery of healthcare services and the ability of professionals to meet public health needs. Institutionalism focuses on the role of institutions in shaping policy and practice within the healthcare system ^10^. It explores how organisational structures, norms, and cultures influence the implementation of regulations. Comparative Policy Analysis assesses policy differences and their impacts across jurisdictions ^3^. By comparing different legal frameworks, it identifies best practices and potential areas for reform. Health Crisis Management considers the effectiveness of regulations in responding to emergencies and crises ^11^. It examines the adaptability of legal frameworks during public health emergencies, such as pandemics or natural disasters. Each framework provides a unique lens for understanding the challenges faced by paramedics when using unapproved medicines and the implications for patient care.

### Data

The following primary legal texts were examined. New Zealand: *Medicines Act 1981* (Section 29) and *Medicines Regulations 1984* ^12, 13^; Australia: *Therapeutic Goods Act 1989* (Special Access Scheme) ^4^; United Kingdom: *Human Medicines Regulations 2012* (Regulation 174) ^5^. These documents were sourced from official government websites and legislative archives. Additionally, the Therapeutic Products Act 2023 in New Zealand was considered ^14^. Additional data were drawn from operational protocols for paramedics, including clinical practice guidelines from ambulance services in each country.

### Comparative Framework

The analysis focused on several key aspects to comprehensively evaluate the regulatory frameworks. Firstly, it examined prescribing and administration authority by investigating who is authorised to prescribe or administer unapproved medicines in each jurisdiction and how this affects paramedics’ ability to act in emergencies. Secondly, the study analysed the administrative requirements by evaluating the notification and reporting obligations imposed on healthcare professionals. This aspect aimed to understand how these obligations impact paramedics’ ability to deliver timely care, considering that extensive administrative processes can hinder prompt medical intervention. Thirdly, the flexibility in emergency situations was assessed by considering how legal frameworks adapt to the demands of emergency medical care. The analysis investigated how regulations accommodate the need for rapid decision-making, which is essential for effective pre-hospital care where delays can have severe consequences. Lastly, the study evaluated the impact on patient care by focusing on how legal frameworks affect paramedics’ ability to provide life-saving interventions. This included examining patient outcomes, and the quality of care delivered under different regulatory conditions, highlighting the practical implications of the laws on the effectiveness of emergency medical services.

### Ethical Analysis

Ethical considerations were explored, particularly the balance between regulatory oversight and the need for immediate decision-making by paramedics. The potential consequences of delaying care or administering unapproved medicines without legal authorisation were examined within the context of patient safety, professional accountability, and the ethical obligations of healthcare providers ^15^.

## Results

A summary of findings is outlined in **Table 1.**

**Table 1.** Summary Table of Findings.

| **Aspect** | **New Zealand**<br>**(Section 29)** | **UK (Human Medicines**<br>**Regulations 2012)** | **Australia (Therapeutic**<br>**Goods Act 1989)** | **New Zealand (Therapeutic**<br>**Products Act 2023)** |
| --- | --- | --- | --- | --- |
| Reporting Requirements | Comprehensive and immediate reporting to the Director-General required | Streamlined post-event reporting for emergency use | Embedded in health system protocols, allowing retrospective reporting | Allows reporting exemptions in emergencies under specific declarations by the Chief Executive |
| Scope for Paramedics | Limited to delegated authority with detailed oversight | Authorised to administer under NHS-approved protocols | Permitted under state-based authorisations through the TGA | Provisions for emergency exemptions could enable paramedics to act more independently but remain untested for implementation |
| Emergency Use Flexibility | High control; delays real-time emergency interventions | Allows temporary use under structured guidelines | Special access schemes enable rapid response | Emergency declarations under Section 116 allow otherwise restricted activities but require Ministerial oversight |
| Administrative Burden | High. Each instance requires pre-approval and detailed reporting, | Low. Emergency-use provisions simplify administration with predefined protocols, | Medium. Reporting requirements exist but are retrospective and aligned with broader | Moderate. Emergency declarations simplify requirements but still rely on higher-level administrative action for |

| Aspect | New Zealand<br>(Section 29) | UK (Human Medicines<br>Regulations 2012) | Australia (Therapeutic<br>Goods Act 1989) | New Zealand (Therapeutic<br>Products Act 2023) |
| --- | --- | --- | --- | --- |
|  | delaying timely interventions and increasing workload. | reducing paramedics' documentation burden. | state and federal systems. | flexibility. |
| Patient Safety vs. Practicality | High emphasis on safety but may delay treatment | Balanced approach supporting rapid action and oversight | Supports timely intervention with safety protocols | Emphasis on safety, but flexibility depends on activating emergency provisions, potentially limiting responsiveness. |
| Health System Integration | Less integrated; high administrative burden | Integrated into NHS frameworks | Integrated into broader health system with state and federal support | Promises improved integration but lacks specific inclusion of paramedics, particularly critical care roles. |

### New Zealand’s Section 29: Comprehensive but Rigid

Section 29 of the *Medicines Act 1981* restricts the use of unapproved medicines to registered medical practitioners and requires individual case notifications to the Ministry of Health ^12^. This framework presents significant challenges for paramedics. In terms of time-sensitive care, the requirement for individual notifications is impractical during emergencies where immediate action is necessary. Paramedics operate in high-pressure environments, and delays in administering treatment can have life-threatening consequences. The administrative process mandated by Section 29 does not accommodate the urgency required in emergency medical situations.

The restricted authority under Section 29 means that paramedics are not authorised to administer unapproved medicines independently, leading to delays when medical practitioners are unavailable. This limitation is particularly problematic in rural or remote areas where access to registered medical practitioners is limited. Paramedics, often the first and only responders, are hindered by legal constraints that prevent them from providing essential care promptly.

The administrative burden of reporting obligations adds to paramedics’ workload, detracting from patient care. In emergency situations, paramedics must prioritise patient stabilisation and rapid decision-making, making extensive documentation impractical. The requirement to report each instance of unapproved medicine use imposes an unrealistic expectation on paramedics during critical moments.

These constraints hinder paramedics’ ability to provide timely interventions, forcing them to choose between delaying treatment or risking legal repercussions. As a result, patient outcomes may be adversely affected, and paramedics face ethical dilemmas in fulfilling their duty of care. The rigidity of Section 29 does not align with the practical necessities of pre-hospital emergency care.

### Equity Considerations for Rural Paramedics

Rigid frameworks like Section 29 disproportionately affect rural paramedics and residents, where healthcare access is already limited due to shortages of medical practitioners. These restrictions lead to delays in patient care and place an additional burden on paramedics in isolated areas who are unable to access support from registered medical practitioners in a timely manner. This is not only a patient care issue but also an equity issue, as rural residents are disproportionately impacted by the inability to provide timely interventions. Flexible frameworks like those in the UK and Australia mitigate these disparities by authorising paramedics to act under emergency protocols, reducing reliance on medical practitioners in rural settings.

A recent study (REF) highlights the disparity in health outcomes for rural residents in New Zealand compared to urban areas, further reinforcing the need for adaptable regulations that support paramedics in these settings.

### Australia: Special Access Scheme (SAS)

Australia’s *Therapeutic Goods Act 1989* provides a more flexible approach through the Special Access Scheme (SAS). Under Category A, healthcare practitioners, including paramedics, can administer unapproved medicines without prior approval for patients with life-threatening conditions, with post-event notification to the Therapeutic Goods Administration (TGA). This provision acknowledges the urgency of emergency care and reduces delays associated with obtaining prior authorisation.

The emergency flexibility of the SAS recognises the realities of pre-hospital care, enabling prompt action while maintaining accountability ^16^. By allowing post-event reporting, the administrative burden does not impede immediate patient care. Paramedics can focus on delivering essential interventions without the hindrance of time-consuming documentation during emergencies.

This framework empowers paramedics to act in the best interests of their patients without being hindered by administrative processes. It strikes a balance between regulatory oversight and practical necessity, ensuring patient safety and effective care delivery. The SAS supports paramedics’ professional judgment and enhances their ability to respond effectively in critical situations.

### United Kingdom: Regulation 174

The UK’s *Human Medicines Regulations 2012* (Regulation 174) offers a streamlined approach. Temporary authorisations permit unapproved medicines during emergencies, following a risk-benefit analysis by the Secretary of State. This mechanism allows for rapid response to public health crises or when approved alternatives are unavailable, facilitating quick adaptation to emerging health needs.

The establishment of national guidelines enables paramedics to administer unapproved medicines under predefined protocols, ensuring legal support without complex approvals. This system provides clarity and confidence for paramedics to make critical decisions. It ensures that paramedics are legally protected when acting within established guidelines, promoting consistency in emergency care practices.

This framework supports paramedics in delivering timely care and enhances the overall responsiveness of the healthcare system. By maintaining regulatory oversight while allowing for flexibility in emergencies, Regulation 174 ensures that patient safety is upheld without compromising the effectiveness of emergency medical interventions.

## Discussion

### The Incompatibility of Section 29 with Pre-Hospital Emergency Care

Section 29’s rigid requirements are incompatible with the urgent nature of pre-hospital emergency care. The restrictions on paramedic authority and the administrative burdens impede timely interventions, potentially compromising patient outcomes. Paramedics are often the sole healthcare providers on the scene and need the autonomy to make immediate decisions without unnecessary legal constraints.

The restricted authority under Section 29 means that paramedics are not authorised to administer unapproved medicines independently, leading to delays when medical practitioners are unavailable. This limitation is particularly problematic in rural or remote areas where access to registered medical practitioners is limited ^17^. Paramedics, often the first and only responders in these regions, are hindered by legal constraints that prevent them from providing essential care promptly. This situation exacerbates health disparities between urban and rural populations, raising equity concerns. Rural residents may face longer response times and reduced access to timely interventions, potentially leading to poorer health outcomes compared to their urban counterparts ^14,17^. Addressing these legal barriers is essential not only for improving patient care but also for promoting health equity across different geographic regions.

The administrative demands of individual case notifications are impractical in emergency settings. Paramedics must focus on patient stabilisation and cannot afford to engage in time-consuming documentation during critical moments. This situation creates ethical dilemmas, forcing paramedics to choose between adhering to the law and providing optimal patient care.

### Comparative Insights: Lessons from Australia and the UK

Australia’s SAS and the UK’s Regulation 174 demonstrate how legal frameworks can balance flexibility with oversight. By allowing paramedics to administer unapproved medicines with post-event reporting, these countries support timely care while ensuring accountability ^4, 5^. These models recognise the unique challenges of emergency medical services and provide practical solutions.

The flexibility in these frameworks empowers paramedics to act decisively, improving patient outcomes. The emphasis on post-event reporting reduces administrative burdens during emergencies without compromising regulatory control. These approaches align legal requirements with the realities of pre-hospital care.

### Ethical Considerations

Delaying care due to legal constraints raises significant ethical concerns. Paramedics have an obligation to provide the best possible care, and restrictive laws like Section 29 may force them to choose between legal compliance and patient welfare ^15^. This conflict can undermine professional integrity and negatively impact patient trust. Flexible frameworks in Australia and the UK better align with ethical imperatives by empowering paramedics to act swiftly in emergencies ^16^. They support the ethical principle of beneficence, enabling healthcare providers to act in the patient’s best interest while maintaining appropriate oversight ^16^.

### Regulatory Theory and Public Health Law

From a regulatory theory perspective, Section 29 represents a high-control model that may be too rigid for emergency care ^6, 8^. It fails to accommodate the need for flexibility in dynamic situations. The comparative analysis suggests that regulations should be adaptable to different contexts, especially in critical care environments. Public health law advocates for regulations that do not unduly hinder access to care, emphasising the need for proportionality ^9, 18^. Laws should protect public health without imposing unnecessary barriers to essential services. Reforming Section 29 to allow greater paramedic autonomy would align with these principles.

### Institutionalism and Policy Implications

Institutions play a crucial role in shaping effective policies. In New Zealand, the Ministry of Health could collaborate with paramedic organisations to reform Section 29, drawing on successful models from Australia and the UK ^10^. Engaging stakeholders ensures that policies are grounded in practical experience and meet the needs of those on the front lines. Implementing changes requires institutional support and a willingness to adapt existing structures. By fostering a collaborative approach, reforms can be more effectively integrated into the healthcare system, enhancing overall efficiency and responsiveness.

### Health Crisis Management: The Need for Adaptable Regulations

The COVID-19 pandemic highlighted the necessity for adaptable regulations. Flexible legal frameworks enable healthcare systems to respond effectively to crises, ensuring continuity of care ^11, 19^. Rigid laws like Section 29 hinder the ability to address unexpected challenges, potentially exacerbating public health emergencies. Adopting more flexible regulations supports resilience in the healthcare system. It allows for rapid deployment of resources and empowers healthcare professionals to act decisively when faced with novel threats.

### Would the Therapeutic Products Act 2023 have addressed these issues for New Zealand?

The Therapeutic Products Act 2023 introduced a framework that appeared to address some of the limitations of Section 29, particularly with respect to emergency-use provisions. However, while this Act is in the process of being repealed, its clauses offer insight into potential future directions for New Zealand’s regulatory landscape ^20^. Key provisions of the Act include:

1. Revisions to Market Authorisation: The bill allows certain activities involving unauthorised therapeutic products under controlled conditions ^20^. In general, these were outlined in Section 9 of the Act. This could potentially have provided more flexibility compared to Section 29 of the Medicines Act 1981.
2. Emergency Arrangements: Section 119 of the Therapeutics Product Act 2023 mentions that in emergency situations, the chief executive of the Ministry could make notices that allow otherwise restricted activities, which would potentially benefit emergency services like paramedics by permitting them to use unapproved medicines in urgent situations without immediate compliance hurdles. Looking at the Therapeutic Products Bill (which contains information not in the Act), the Regulations Review Committee suggested that these emergency powers should not be delegated by the minister to the Chief Executive, quite a contrast to both the Australian and United Kingdom approaches ^21^.
3. Special Case Provisions: Sections 66 and 89 discuss specific cases where health practitioners can import and supply unapproved medicines under conditions that satisfy regulatory requirements, providing more leeway than the current Section 29.

These updates appear as though they may have addressed some of the administrative and regulatory constraints that paramedics currently face, by potentially simplifying the use of unapproved medicines during emergencies and enabling a more responsive approach. However, while these changes might offer improvements, a future bill should address how common and serious scenarios would be addressed in practice, and that real-time administration without excessive delays or post-event penalties is feasible.

As a final point with regards to the TPA, it is notable that although nurse practitioners are mentioned, specialist paramedics are not despite being a substantially similar model and indeed the critical care paramedical nature of the role having outgrown the delegated standing orders system, warrants a regulatory approach which is more fit for purpose and the actual role that paramedics play in New Zaland society.

### Recommendations for New Zealand’s Framework

A summary of recommendations is available in **Table 2**. To address the challenges identified in the current regulatory approach, several key recommendations are proposed to reform New Zealand’s framework concerning the use of unapproved medicines by paramedics. Firstly, expanding paramedic authority is crucial. Amending a future medicines bill to include paramedics would allow them to administer unapproved medicines under defined conditions-such as a scope of clinical practice already regulated by the *Health Practitioners Competence Assurance Act (2003)* system. This change would acknowledge their expertise and critical role in emergency care, empowering them to act promptly in life-threatening situations without unnecessary legal constraints but whilst reasonably expecting similar public safety oversight such as medical practitioners, veterinarians, dentists and nurse practitioners. Secondly, there is a need to simplify administrative requirements. Introducing post-event reporting would reduce administrative burdens during emergencies. By streamlining documentation processes, accountability is maintained without hindering immediate action. This approach ensures that paramedics can focus on delivering timely patient care during critical moments and meaning that regulatory processes do not directly hinder appropriate life-saving clinical action.

**Table 2.** Summary of Recommendations.

| **Recommendation** | **Justification** | **Expected Outcome** | **Addressed by TPA?** |
| --- | --- | --- | --- |
| Controlled Authorisation | Aligns with practices in the UK and Australia | Expanded paramedic scope, timely care | Partially; TPA allowed certain activities under controlled conditions but did not explicitly expand paramedic scope |
| Emergency Exemption Amendment | Reduces delay while maintaining post-event oversight | Faster emergency response, compliance | Yes; TPA included emergency provisions for the chief executive to permit restricted activities |
| Tiered Training and Certification | Ensures safety with expanded prescribing rights | High level of trust and patient safety | Not specifically addressed; TPA did not outline training programs for paramedics |
| Integrated Reporting Systems | Simplifies compliance without real-time delays | Efficient reporting, reduced admin burden | Partially; TPA proposed more flexible reporting requirements |
| Collaborative Policy Development | Strengthens policy through stakeholder input | Enhanced policy acceptance and effectiveness | Uncertain; TPA development may not have fully engaged all stakeholders, including paramedic associations |

Thirdly, establishing emergency use provisions would provide clear legal support. Creating protocols for the use of unapproved medicines in emergencies, like Australia’s Special Access Scheme and the UK’s Regulation 174, would offer consistency in practice. Clear clinical practice guidelines are already available to enable paramedics to make critical decisions confidently, but what is necessary is for them and the medical practitioners supporting them to know they are operating within a legally supported framework. Additionally, it is recommended to develop tiered training programs to certify paramedics for administering unapproved medicines. Implementing specialised education ensures competence and safety, reinforcing professional standards and building patient trust. Such training would equip paramedics with the necessary skills and knowledge to handle unapproved medicines appropriately. In the current environment where ambulance services are privately operated under a partly publicly funded arrangement, there are not resources available for that to be organisationally supported.

Finally, fostering institutional collaboration is essential for effective policy development. Engaging stakeholders—including paramedic associations, healthcare institutions, and policymakers—in the reform process ensures that new regulations meet practical needs. We often hear the expression from public health circles, “The Ambulance at the bottom of the cliff”. This ignores the substantial growth of paramedics and ambulance services in providing both core emergency medical services but the 30+ year expansion into urgent care, and the fact that they are now the ambulance well before, at the top, half-way down, as well as the bottom of the cliff. That preventable acute conditions prevent via ambulance services, does not unfortunately mean that an austere funding approach improves health outcomes. Collaborative efforts enhance the relevance and effectiveness of the regulations, promoting widespread acceptance and successful implementation. By adopting these recommendations, New Zealand can create a more flexible and responsive regulatory framework that empowers paramedics, maintains patient safety, and aligns with international best practices. These changes would ultimately improve patient outcomes during emergencies and strengthen the healthcare system’s ability to respond effectively to critical situations.

## Conclusion

New Zealand’s Section 29 of the *Medicines Act 1981* is not fit for purpose in pre-hospital emergency care. The legal restrictions and administrative burdens hinder paramedics’ ability to provide timely, life-saving interventions. This situation poses ethical dilemmas and potentially compromises patient outcomes.

In contrast, Australia and the UK offer flexible frameworks that empower paramedics while maintaining oversight. These models demonstrate that it is possible to balance regulatory control with practical necessity, supporting both patient safety and effective care delivery.

Urgent legal reform is needed in New Zealand to expand paramedic authority, simplify administrative processes, and establish emergency use provisions. By aligning with international best practices, New Zealand can enhance paramedic autonomy, improve patient outcomes, and ensure that regulatory frameworks support effective emergency responses.

Such changes would prevent paramedics from facing legal risks or compromising patient care, ultimately strengthening the healthcare system’s ability to respond to emergencies. Embracing flexibility and collaboration in regulatory practices is essential for meeting the evolving challenges of modern healthcare.

## Data Availability

All data produced are available online.

## Acknowledgements

Nil

## Ethics

Nil

## Data

The legislation reviewed in this research is all outlined in the references and publicly available.

## References

1. Ivanov D. Predicting the impacts of epidemic outbreaks on global supply chains: A simulation-based analysis on the coronavirus outbreak (COVID-19/SARS-CoV-2) case. Transportation Research Part E: Logistics and Transportation Review. 2020; 136:101922.

2. Kavanagh MM, Singh R. Democracy, Capacity, and Coercion in Pandemic Response: COVID-19 in Comparative Political Perspective. J Health Polit Policy Law. 2020; 45:997–1012.

3. Blank RH, Burau V, Kuhlmann E. Comparative Health Policy: Macmillan Education UK, 2017.

4. Therapeutic Goods Act 1989 Australia, 1989.

5. Human Medicines Regulations 2012. 2012.

6. Baldwin R, Cave M, Lodge M. Understanding Regulation: Theory, Strategy, and Practice: OUP Oxford, 2012.

7. United Nations Office for Disaster Risk Reduction. The human cost of disasters: an overview of the last 20 years (2000-2019). United Nations Office for Disaster Risk Reduction, 2021.

8. De Cruz P. Comparative healthcare law: Routledge, 2013.

9. Gostin LO, Wiley LF. Public Health Law: Power, Duty, Restraint: University of California Press, 2016.

10. Hall PA, Taylor RCR. Political Science and the Three New Institutionalisms. Political Studies. 1996; 44:936–57.

11. Ansell C, Boin A, Keller A. Managing Transboundary Crises: Identifying the Building Blocks of an Effective Response System. Journal of Contingencies and Crisis Management. 2010; 18:195–207.

12. Medicines Act 1981. New Zealand, 1981.

13. Medicines Regulations 1984. New Zealand, 1984.

14. Te Tāhū Hauora Health Quality & Safety Commission. Te kounga o te tauwhiro hauora Window on the quality of health care. Wellington, New Zealand: Te Tāhū Hauora Health Quality & Safety Commission, 2021.

15. Beauchamp TL, Childress JF. Principles of Biomedical Ethics: Oxford University Press, 2001.

16. O’Meara P, Wingrove G, Nolan M. Clinical leadership in paramedic services: A narrative synthesis. International Journal of Health Governance. 2017; 22:251–68.

17. Ministry of Health. Health and Independence Report 2022 Te Pūrongo mō te Hauora me te Tū Motuhake 2022. Wellington: Ministry of Health, 2022.

18. Gostin LO. Pandemic Influenza: public health preparedness for the next global health emergency. J Law Med Ethics. 2004; 32:565–73.

19. Legido-Quigley H, Mateos-García JT, Campos VR, Gea-Sánchez M, Muntaner C, McKee M. The resilience of the Spanish health system against the COVID-19 pandemic. Lancet Public Health. 2020; 5:e251–e2.

20. Therapeutic Products Act New Zealand, 2023.

21. Therapeutic Products Bill 2022.

