## Supplementary Document for "Unapproved Medicine Use by Paramedics in New Zealand: A Comparative Analysis with Australian and UK Frameworks"

**Supplementary Tables**

**Framework-Based Analysis of Medicine Regulation Across Jurisdictions**

| **Framework** | **UK** | **Australia** | **New Zealand** |
| --- | --- | --- | --- |
| Legal Pluralism | Multiple layers of regulation; Medicines Act 1968 and Human Medicines Regulations blend national and international influences. | Federal oversight by TGA, with integration of state regulations. | Combines domestic law with global practices, evident in Medicines Act 1981. |
| Regulatory Theory | Strict regulation under the Medicines Act 1968; emergency provisions in Human Medicines Regulations 2012. | The Therapeutic Goods Act 1989 governs, with emergency access through TGA's special access schemes. | Section 29 allows unapproved medicines with strict reporting to the Director-General. |
| Comparative Legal Analysis | Strong alignment with EU directives pre-Brexit; post-Brexit shifts expected. | Reflects federal-state legal integration and alignment with international standards. | Clear distinctions in regulations for handling unapproved medicines under Section 29. |
| Public Health Law Framework | Emphasises public safety and controlled access to unapproved medicines. | TGA ensures public health and regulated emergency access to unapproved medicines. | Focuses on emergency supply with oversight to ensure public health safety. |
| Institutionalism | Institutions like MHRA enforce strict compliance and oversight. | TGA oversees regulations and enforcement, supporting institutional adherence. | Ministry of Health ensures adherence through Director-General oversight. |
| International Health Law Framework | Historically influenced by EU, ensuring international alignment pre-Brexit. | Aligns with WHO and global standards in therapeutic goods regulation. | Aligned with global practices for emergency medicine use while maintaining local control. |
| Sociological Legal Framework | Considers societal safety and controlled use of medicines in emergencies. | Focuses on safe use and accessibility of emergency medicines. | Emphasises oversight and societal impacts, especially in emergencies. |
| Emergency Medical Systems Framework | Emergency use defined in regulations for controlled temporary authorisations. | Allows for emergency and compassionate use with TGA oversight. | Section 29 and Medicines Regulations 1984 specify emergency protocols. |
| Health Crisis and Disaster Law | Supports the use of unapproved medicines in public health emergencies. | Supports rapid response with emergency amendments for unapproved drugs. | Handles supply chain issues by permitting emergency use under strict reporting. |
| Scope of Practice and Professional Autonomy | Paramedics require NHS guidance and specific authorisations for use. | State laws dictate paramedic authority for emergency medicine use. | Limited prescribing authority for paramedics under delegation and protocols. |
| Pharmaceutical Regulation and Access Framework | Robust processes under Medicines Act 1968 and Human Medicines Regulations 2012. | TGA's framework supports regulation and controlled access to unapproved drugs. | Section 29 outlines Ministerial consent for unapproved medicine use. |
| Health Workforce and Role Delineation | Roles defined under NHS guidance; strict authorisation for paramedics. | Roles defined by state regulations; paramedics need authorisation for emergency administration. | Defines paramedic roles under delegated authority for emergency situations. |
| Comparative Policy Analysis | Detailed regulatory approach emphasising controlled and documented use. | Adaptive approach for emergency and urgent needs under TGA schemes. | Specific Section 29 and emergency provisions are key for handling supply issues. |
| Health Equity and Access to Care | Ensures equitable access to unapproved medicines in emergencies. | Provisions support equity in access during medical emergencies. | Ensures continuity of care during supply chain disruptions with oversight. |

**Comparison of Provisions for Unapproved Medicines Across Jurisdictions**

| **Jurisdiction** | **Regulatory Framework** | **Ease of Use in Emergencies** | **Adequacy for Paramedics** |
| --- | --- | --- | --- |
| UK | Human Medicines Regulations 2012 - Temporary authorisation for emergency use. | Flexible, allows paramedics to act quickly within set guidelines. | Supportive, reduces on-the-spot reporting and administrative burden. |
| Australia | Therapeutic Goods Act 1989 - Special Access Schemes for unapproved medicines. | Adaptable for pre-hospital care with state-level authorisations. | Practical, balances regulation with emergency care flexibility. |
| New Zealand | Medicines Act 1981 - Section 29 mandates strict reporting for unapproved use. | Restrictive; extensive reporting hinders timely use in emergencies. | Impractical; paramedics may avoid use due to stringent reporting. |

**Practical Options for Administering Unapproved Medicines in New Zealand**

| **Option** | **Description** | **Challenges** |
| --- | --- | --- |
| Protocol-Based Pre-Authorisations | Implement standing orders or pre-approved protocols under medical oversight. | May require legislative changes for broader protocol acceptance. |
| Delegated Medical Authority | Remote supervision by an on-call medical director for reporting responsibility. | Real-time communication challenges in rural or busy environments. |
| Emergency Exemption Clauses | Amend Section 29 to allow specific emergency exemptions. | Legislative amendments needed; safeguards required to prevent misuse. |
| Streamlined Reporting System | Develop digital tools for real-time or shift-end reporting. | Adds administrative steps but is less burdensome than current requirements. |
